# Arm-specific versus total-sample fragility quotients in large vessel vasculitis trials

**DOI:** 10.64898/2025.12.01.25341404

**Authors:** Justin J. Gillette, Michelle Lu, Daniel Y. Tsybulnik, Thomas F. Heston

## Abstract

The fragility quotient normalizes the fragility index by total sample size but can obscure arm-specific fragility when trial arms are unequal, since the fragility index systematically alters outcomes in only one arm. We defined the modified-arm fragility quotient as the fragility index divided by the size of the arm in which outcomes are toggled. We compared it to the fragility quotient using three approaches: (1) re-analysis of 29 categorical outcomes from 18 large-vessel vasculitis trials (allocation ratios 1.0–2.0), (2) R grid simulation across 1:1 to 4:1 allocation ratios with sample sizes 60–240 (90,000 simulations), and (3) Python Monte Carlo simulation with 3,000 replications across 1:1, 1:2, and 1:3 allocations. In balanced 1:1 trials, the modified-arm fragility quotient equaled twice the fragility quotient exactly, as predicted. The proportion of trials in which the modified-arm fragility quotient exceeded twice the fragility quotient increased systematically with allocation imbalance: 0% (1:1), 81% (2:1), 93% (3:1), and 95% (4:1). In the R simulation, mean divergence between the modified-arm fragility quotient and twice the value of the fragility quotient increased from 0.00 (1:1) to 0.10 (4:1). Empirical data showed a strong correlation (r = 0.98) between the two quotients, but divergence increased in the 25 unbalanced allocation trials. The ratio of the modified-arm fragility quotient to twice the value of the fragility quotient scaled linearly with allocation ratio, with mean values of 1.0 (1:1), 1.36 (2:1), 1.91 (3:1), and 2.41 (4:1). These findings show that the modified-arm fragility quotient and total-sample fragility quotient convey equivalent information under equal allocation. In contrast, under unequal randomization, the total-sample fragility quotient can mischaracterize trial fragility due to dilution from the larger, unmodified arm, whereas the modified-arm fragility quotient avoids this bias by restricting normalization to the modified arm.

## Introduction

Large vessel vasculitis (LVV) refers to inflammation of the aorta and its major branches and is broadly categorized into giant cell arteritis and Takayasu arteritis [1]. Glucocorticoids have been the mainstay of treatment for LVV, but other agents like interleukin-6 inhibitors and disease-modifying antirheumatic drugs (DMARDS) have been investigated to reduce glucocorticoid dependence [2]. Prolonged glucocorticoid therapy is associated with an increased risk of diabetes, infections, and cardiovascular disease, with studies reporting that up to 86% of patients develop glucocorticoid-related adverse effects during follow-up [3]. These high rates of glucocorticoid-related complications highlight the urgent need for well-designed clinical trials to evaluate new therapeutics and optimize treatment strategies for patients with LVV. As trials for the treatment of LVV increase, the need for fragility metrics becomes essential.

Randomized controlled trials (RCTs) often rely on a dichotomous threshold of statistical significance (e.g., p < 0.05) to guide clinical decision-making. Yet this approach does not capture how easily a result could change if only a few outcomes were different [4]. The fragility index (FI) quantifies the minimum number of outcome reversals needed to change statistical significance [5]. To reduce its dependence on sample size, the fragility quotient (FQ) divides FI by total enrollment (N), improving cross-trial comparability [6]. However, because FI toggles occur within a single arm, dividing by N can misstate arm-level fragility under unequal allocation. Unequal allocation is increasingly common in rare disease research, where enrollment constraints or ethical considerations favor assigning more participants to active treatment [7]. Small populations, low event rates, and asymmetric randomization combine to test the limits of conventional fragility metrics [8].

Although FQ normalizes FI by N, it inherits FI’s structural limitations: it is tied to a binary threshold (the p-value), and does not incorporate the probability of outcome modifications [9], and depends on the chosen definition of an event. FQ also uses a total-sample denominator even though only one arm drives FI; this can overstate fragility when the modified arm is smaller. FQ may misrepresent arm-specific fragility when allocation ratios deviate from 1:1, because the denominator blends both study arms and thus becomes sensitive to the allocation ratio [10]. For example, in a trial of 150 patients, with 100 in the treatment arm and 50 in the control arm, if the FI is 5, the FQ would be 5/150 = 0.033. Although this number suggests a moderately fragile result, changing the results for 5 out of 50 patients in the control arm would be 10% of that group, suggesting that these results are more stable than the FQ indicates.

To address these issues, we evaluate the modified-arm fragility quotient (MFQ), defined as FI divided by the size of the arm receiving the outcome toggles. In balanced 1:1 designs, MFQ and FQ convey equivalent information (MFQ/2 = FQ). However, under unequal allocation, they will diverge systematically: MFQ preserves arm-specific meaning, whereas FQ may overstate fragility by yielding an artificially low value due to total-sample scaling. Using re-analysed categorical outcomes from 29 large vessel vasculitis trials, a large-scale grid simulation in R across allocation ratios 1:1 to 4:1 with sample sizes 60–240, and a Python Monte Carlo simulation reflecting realistic trial conditions, we identify when FQ suffices and when MFQ is required to avoid overstatement of trial fragility.

Rare disease trials, including those in large vessel vasculitis (LVV), frequently employ unequal allocation due to ethical constraints, adaptive designs, or multi-arm comparisons [11,12]. We empirically evaluated MFQ using data from a systematic review of LVV trials to demonstrate its utility in assessing arm-specific fragility. While FI and FQ have previously been investigated in LVV trials [13,14], no studies have evaluated whether applying MFQ alters the interpretation of trial fragility. Since unequal allocation is becoming more common in rare disease trials, using MFQ could provide a more clinically meaningful way to assess result stability by evaluating the fragility of the arm that contributed to statistical significance.

## Methods

### Data sources

The empirical component re-evaluated 29 categorical outcomes from 18 randomized controlled trials reported by Misra et al.[13]. No new literature search was performed, nor were raw, unpublished data collected. All primary source manuscripts were previously published online, publicly available, and independently reviewed to verify outcome counts. For each outcome, we computed the baseline two-sided Fisher’s exact p-value, the post-toggle p-value, the fragility index (FI), fragility quotient (FQ), modified-arm fragility quotient (MFQ), the allocation ratio scaling factor ρ = MFQ / (2×FQ), the sizes of the toggled and untoggled arms, and whether the baseline result was statistically significant (p ≤ 0.05).

To generalize beyond this empirical sample, we conducted two complementary simulations. The first simulation was performed using R Statistical Software (v4.5.1; R Core Team 2025) [15]. This R grid simulation employed a factorial design varying total sample size N (60, 120, 240), allocation ratio (1:1, 2:1, 3:1, 4:1), control arm risk (0.05, 0.20, 0.40), and risk ratios (0.75, 0.90, 1.00, 1.10, 1.25), with 500 replicates per parameter combination across 180 distinct conditions, yielding 90,000 total simulations.

The Python simulation (Python version 3.10+) [16] generated 1,000 valid 2×2 contingency tables for each of three fixed allocations: 1:1 (n_1_=50, n_2_=50), 1:2 (n_1_=35, n_2_=70), and 1:3 (n_1_=30, n_2_=90), with event probabilities sampled broadly to reflect realistic trial conditions, yielding 3,000 total simulations. For both simulations, FI, FQ, MFQ, and ρ were computed using the same algorithms as in the empirical analysis.

### Fragility metrics

#### Fragility Index (FI)

The minimum number of binary outcome toggles required to cross the p = 0.05 threshold using two-sided Fisher’s exact test. Outcome toggles were applied to the arm with fewer events; if event counts were equal, toggles were applied to the smaller arm. This rule was applied identically to trials with baseline p ≤ 0.05 (counting toggles to reach p > 0.05) and trials with baseline p > 0.05 (counting toggles to reach p ≤ 0.05).

#### Fragility Quotient (FQ)

FI divided by total sample size (N).

#### Modified-arm Fragility Quotient (MFQ)

FI divided by the size of the modified arm (n_mod), where n_mod is the arm receiving outcome toggles during FI calculation. Under a balanced 1:1 allocation, MFQ/2 = FQ by mathematical identity; under an unequal allocation, the two metrics diverge.

#### Allocation ratio scaling factor (ρ)

ρ = MFQ / (2×FQ). Under 1:1 allocation, ρ = 1. Values >1 indicate that FQ overstates fragility relative to MFQ (total-N scaling yields an artificially low FQ when the modified arm is the smaller arm). Values < 1 indicate that FQ understates fragility relative to the MFQ (the modified arm is larger than the opposite arm).

### Statistics

All analyses were performed using SPSS (IBM SPSS Statistics version 31), R (version 4.5.1), Python (version 3.10), and Excel (version 2510, Microsoft Corporation). Distributions were summarized using medians and interquartile ranges. Divergence between FQ and MFQ was quantified as |MFQ/2 − FQ| / FQ, expressed as a percentage. Pearson correlation coefficients were used to assess the linear relationship between FQ and MFQ. Mann-Whitney U tests compared FQ, MFQ, and ρ values between trials with statistically significant versus non-significant baseline results. The agreement between theoretical and observed values of ρ was evaluated using Pearson’s correlation and mean absolute deviation. All hypothesis tests were two-sided with α = 0.05. FI computation assumes valid baseline analyses. No formal bias assessment was performed because the study is methodological, not clinical.

### Reporting standards

This study conformed to the Statistical Analyses and Methods in the Published Literature (SAMPL) guidelines for statistical reporting [17]. The complete SAMPL checklist is provided in Supplementary File S1.

### Ethics

This research analyzed simulated data and re-analyzed previously published, peer-reviewed aggregate data from randomized controlled trials. No individual-level or identifiable participant data were reviewed or used. Because all data were either synthetically generated or obtained from publicly available sources in aggregate form, this work did not require institutional review board approval or informed consent.

## Results

We analyzed 29 categorical outcomes from 18 randomized controlled trials, including 1,397 participants. Sample sizes ranged from 12 to 321 per trial. Twelve outcomes were statistically significant at baseline, and 17 were not. Allocation ratios ranged from 1:1 to 2:1 (mean 1.3:1).

In the empirical data, the Fragility Quotient (FQ) and Modified Fragility Quotient (MFQ) were highly correlated (Pearson r = 0.981, p < 0.001), indicating they quantify the same construct of fragility. MFQ was modestly larger than FQ by design: the mean MFQ / (2 × FQ) was 1.12 (SD 0.17), consistent with the theoretical identity ρ = N / (2·n_mod). “Fragility did not differ by statistical-significance status. Mann–Whitney tests comparing FQ, MFQ, and MFQ/(2×FQ) between significant and non-significant results showed no detectable differences (all p > 0.16). Fragility was not meaningfully associated with total sample size: correlations between N and all fragility measures were small and statistically non-significant across both Pearson and Spearman analyses (all |r| < 0.40). Table 1 summarizes the empirical fragility metrics stratified by allocation ratio.

**Table 1.**
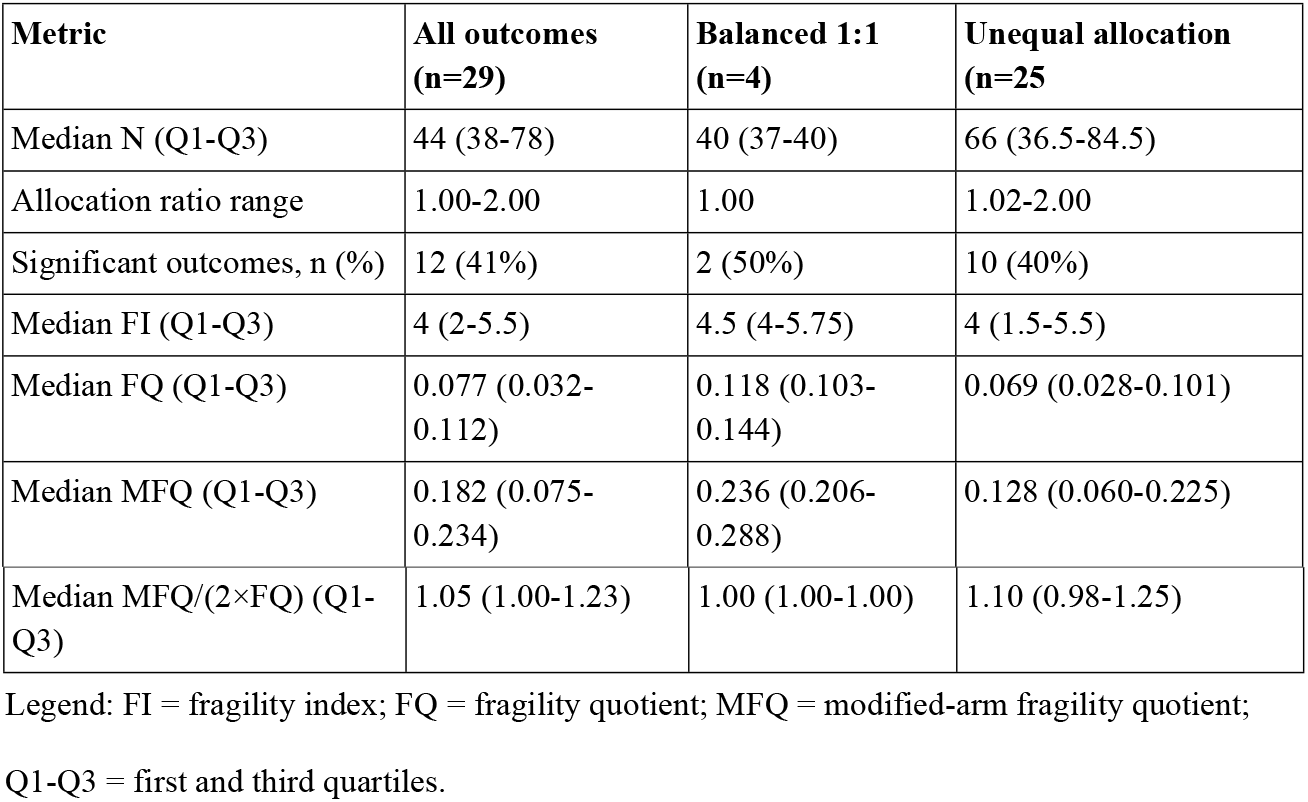
Summary of empirical fragility metrics stratified by allocation ratio.

| Metric | All outcomes<br>(n=29) | Balanced 1:1<br>(n=4) | Unequal allocation<br>(n=25) |
| --- | --- | --- | --- |
| Median N (Q1-Q3) | 44 (38-78) | 40 (37-40) | 66 (36.5-84.5) |
| Allocation ratio range | 1.00-2.00 | 1.00 | 1.02-2.00 |
| Significant outcomes, n (%) | 12 (41%) | 2 (50%) | 10 (40%) |
| Median FI (Q1-Q3) | 4 (2-5.5) | 4.5 (4-5.75) | 4 (1.5-5.5) |
| Median FQ (Q1-Q3) | 0.077 (0.032-0.112) | 0.118 (0.103-0.144) | 0.069 (0.028-0.101) |
| Median MFQ (Q1-Q3) | 0.182 (0.075-0.234) | 0.236 (0.206-0.288) | 0.128 (0.060-0.225) |
| Median MFQ/(2×FQ) (Q1-Q3) | 1.05 (1.00-1.23) | 1.00 (1.00-1.00) | 1.10 (0.98-1.25) |

Legend: FI = fragility index; FQ = fragility quotient; MFQ = modified-arm fragility quotient;
Q1-Q3 = first and third quartiles.

In 18 of 25 unbalanced trials, the smaller arm had fewer events and was toggled when calculating the FI, yielding a mean MFQ/(2×FQ) of 1.21 (SD 0.161). In the remaining 7 trials, the larger arm had fewer events, yielding a mean MFQ/(2×FQ) of 0.954 (SD 0.036). This bidirectional divergence demonstrates that FQ’s fixed total-sample denominator does not track which arm drives fragility, whereas MFQ’s denominator follows the actual toggle pathway.

Both simulation studies reproduced these empirical patterns and reinforced the divergence of the MFQ under unequal allocation. In the R simulation, MFQ and FQ behaved as expected under 1:1 allocation (MFQ = 2 × FQ) and diverged systematically as the imbalance increased. The proportion of replicates with MFQ > (2 × FQ) rose from 0% at 1:1 to 81%, 93%, and 95% at 2:1, 3:1, and 4:1. Mean (SD) MFQ / (2 × FQ) values were 1.00 (0.00), 1.36 (0.29), 1.91 (0.33), and 2.41 (0.40), respectively (Fig 1).

**Figure 1.**
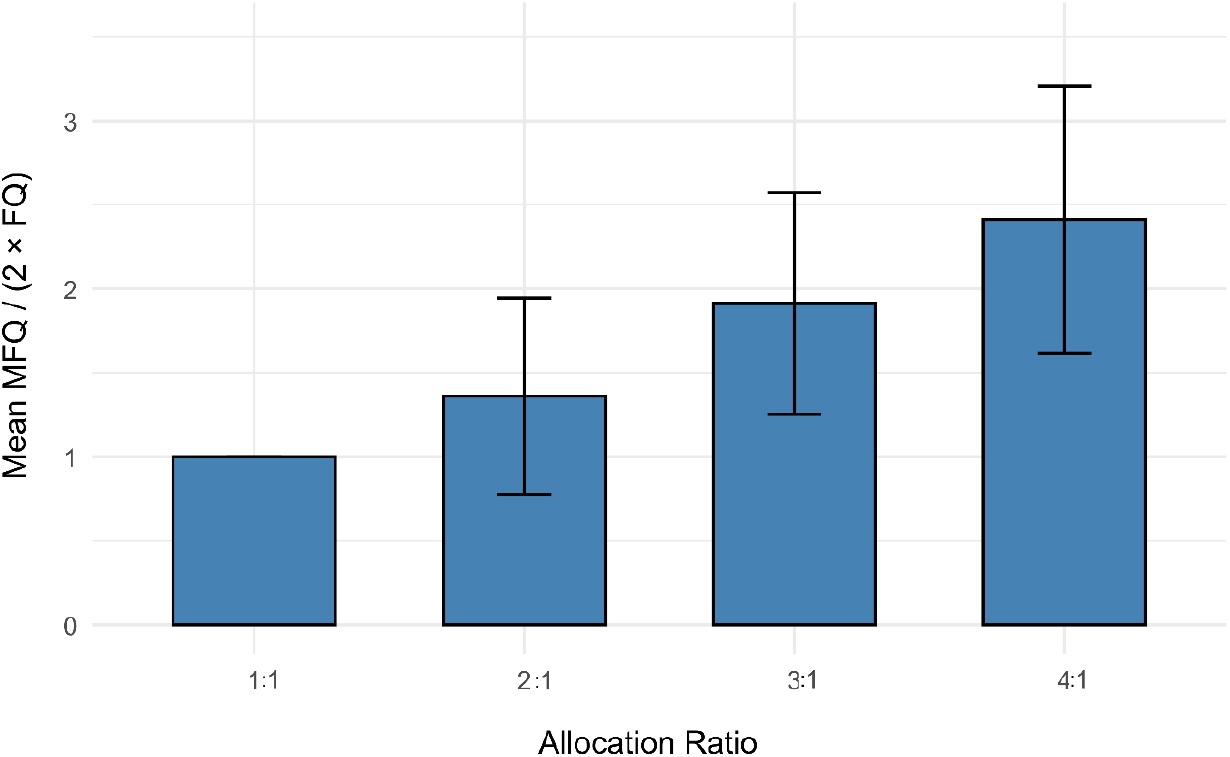
Mean MFQ/(2×FQ) by allocation ratio in R simulation study. Bars show the mean ratio of MFQ to twice the corresponding FQ across 22,500 simulated trials per allocation ratio. Error bars represent ±2 standard deviations. Under balanced allocation (1:1), MFQ equals 2×FQ by design (ratio = 1). As allocation becomes more unequal, MFQ increases systematically relative to FQ, reaching a ratio of 2.41 at 4:1 allocation.

In the Python simulation with fixed arm sizes, MFQ likewise scaled predictably with allocation. The mean (SD) MFQ / (2 × FQ) was 1.00 (0.00), 1.32 (0.32), and 1.85 (0.42) for 1:1, 2:1, and 3:1 allocations, respectively. FQ and MFQ remained strongly correlated at all levels (r = 1.00, 0.89, 0.76; p < 0.001), again indicating that both metrics quantify the same fragility construct, but MFQ appropriately scales by the arm that determines fragility.

Across the three allocation scenarios tested (1:1, 1:2, 1:3), the median FI values were similar (5, 4, and 4, respectively), but the distribution of FI narrowed as allocation became more unequal (Figure 2). Under 1:1 allocation, FI values ranged widely, including a long right tail. Under 1:2 and 1:3 allocation, fewer extreme FI values were observed, and the overall spread compressed despite comparable total sample sizes (100, 105, and 120 participants). Thus, unequal allocation was associated with reduced dispersion of FI, even when absolute FI changed only modestly with total sample size.

**Fig 2.**
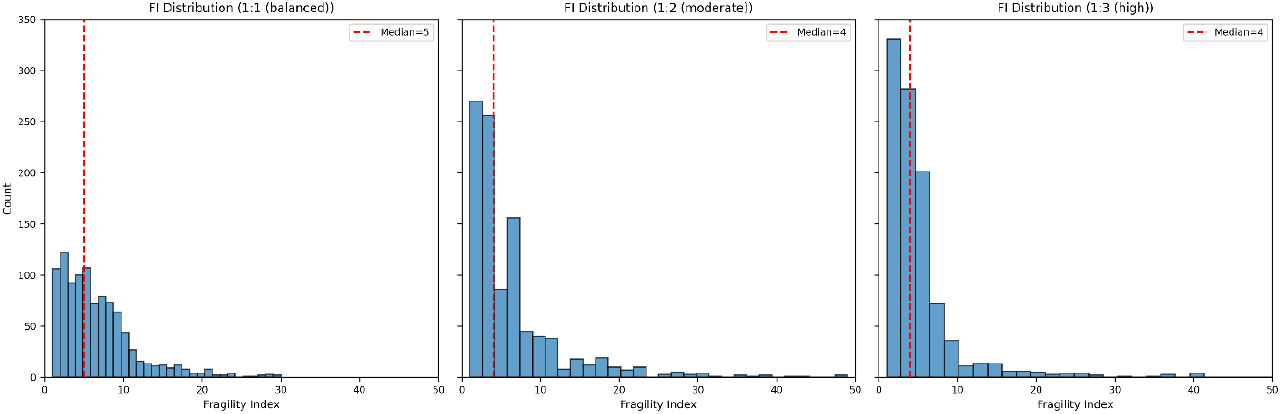
Distribution of FI across allocation ratios in the Python simulation study. Each panel shows FI values from 1,000 simulated 2×2 trials under balanced (1:1), moderate (1:2), and high (1:3) allocation imbalance. Unequal allocation produced visibly narrower FI distributions, reflecting reduced FI variability due to geometric constraints imposed by the smaller toggled arm.

## Discussion

The empirical re-analysis and both simulations converge on a single conclusion: the fragility quotient (FQ) and modified-arm fragility quotient (MFQ) describe the same construct of fragility under balanced allocation, but diverge systematically when randomization is unequal. In 1:1 designs, the two are mathematically equivalent. In imbalanced allocations, the MFQ provides more accurate arm-level scaling because it normalizes the fragility index (FI) to the arm’s sample size where outcome toggles occur. The modest inflation of MFQ over FQ observed in empirical trials (∼12% on average) was fully explained by arm-size ratios and required no additional assumptions. These results support using FQ for balanced trials, but MFQ for unbalanced designs in which arm-specific fragility differs meaningfully from total-sample fragility.

In the empirical dataset, MFQ consistently exceeded FQ in proportion to the smaller arm’s relative size. Both measures were highly correlated, confirming they capture the same underlying quantity. However, in unbalanced randomization, the total-sample denominator in FQ produced artificially low values—overstating fragility—because FI toggles typically occur in the smaller or fewer-events arm. Across the R simulations and Monte Carlo replications, MFQ tracked allocation ratios exactly as predicted by geometry, confirming the relationship is deterministic rather than stochastic.

Beyond the primary aim of comparing FQ and MFQ, the Python simulation also revealed that unequal allocation compresses the FI distribution, reducing both variability and magnitude of fragility. This structural effect likely reflects reduced event flexibility when one arm is larger.

In other specialties, including cardiology, the fragility index has gained popularity as a simple, accessible tool for clinicians to assess the reliability of trial results [18]. However, the FI is inherently sample-size-dependent, which complicates comparisons of fragility across trials of different sizes [19]. While FQ aims to standardize FI by dividing by the population size, it can easily be distorted in RCTs with unequal allocation between study arms. MFQ is a solution, as it normalizes the fragility index to the modified arm rather than the total population. Comparing the MFQ to the FQ enables clinicians to better assess whether an observed treatment effect is stable or depends on outcomes from only a few patients.

When designing clinical trials for new therapies, MFQ can be incorporated into simulations of allocation ratios and event rates to estimate how many outcome reversals in a specific arm would render results significant or non-significant, which could help guide researchers in determining appropriate sample sizes. Tools have already been developed to incorporate the FI into sample size calculations for clinical trials, providing a framework that maintains low Type 1 error rates and offering results that are more intuitive for clinicians to interpret [20]. MFQ would add another level of validity, accounting for unequal allocation ratios and offering insight into which study arm drives fragility. Additionally, in rare disease trials, MFQ could provide a quantitative basis for minimizing patient exposure to inferior treatments while still ensuring interpretable results.

Regulatory agencies such as the Food and Drug Administration (FDA) and the European Medicines Agency (EMA) issue guidance documents outlining key aspects of clinical trials for new drug development, including trial design, trial populations, and statistical considerations [22]. Integrating MFQ into this framework could provide an additional measure of trial stability. Institutional Review Boards (IRBs) may find MFQ helpful when evaluating trial protocols, as it summarizes how many outcome changes in a specific arm would be required to overturn statistical significance. Trials with very low MFQs under planned allocation ratios may warrant re-examination of design features such as sample size or allocation imbalance before exposing patients to novel therapies. Overall, incorporating MFQ into regulatory and ethical oversight processes could increase transparency about the stability of trial findings.

### Future directions

The FI quantifies fragility by iteratively toggling outcomes within a single trial arm, but it remains threshold-dependent and path-specific. Two emerging frameworks address these limitations. First, the Global Fragility Index (GFI) extends fragility measurement to path-independent perturbations, quantifying the minimum total change across all cells of a contingency table required to reverse significance, regardless of which arm or combination of arms contributes to the shift [21]. This approach removes dependence on arbitrary toggle rules and provides a unified fragility measure for complex trial designs beyond two-arm binary outcome studies. The observed narrowing of FI under allocation imbalance highlights a structural limitation of the conventional FI. Future work should evaluate how global measures such as the GFI perform under these same conditions to determine whether global metrics capture fragility more effectively across varying allocation ratios and event distributions.

Second, the Neutrality Boundary Framework (NBF) adds a complementary robustness dimension (nb) that quantifies geometric distance from statistical neutrality, independent of the α = 0.05 threshold [22]. By producing bounded 0–1 robustness scores alongside fragility quotients, also 0-1 bounded, the NBF enables a more complete evidence quality assessment that is directly comparable across outcome types, sample sizes, and study designs. Together, these methods form a coherent system for quantifying statistical significance, fragility, and robustness across the full spectrum of trial architectures.

Suggested practical implications are to: a) incorporate MFQ into trial design to proactively assess how allocation ratios and event distributions affect arm-level fragility; present FI, FQ, and MFQ jointly in trials with unequal randomization, particularly in rare diseases where such designs are common; and b) when MFQ is greater than the FQ, total-sample scaling may have made the FQ artificially low, with the result being that the FQ overstates fragility; the modified arm is less fragile than the total-sample fragility quotient implies.

### Limitations

This study examined categorical outcomes from large vessel vasculitis trials and simulated binary endpoints; we did not assess continuous or time-to-event outcomes. Both FQ and MFQ inherit the fragility index’s dependence on event definition and the α = 0.05 significance threshold, limiting their applicability when continuous fragility measures or threshold-free metrics are required. The empirical dataset was restricted to a single disease domain, and no formal risk-of-bias assessment was performed. However, the core mathematical relationships among arm size, allocation ratio, and fragility scaling generalize to all two-arm binary-outcome trials, and replication of patterns across two independent simulation systems (90,000 R factorial runs and 3,000 Python Monte Carlo replications) strengthens confidence in the findings. All included trials employed pre-specified binary endpoints, and FI computation depends only on valid contingency table data rather than patient-level information. Simulation parameters reflected realistic ranges of risk and allocation, ensuring broad coverage of typical trial conditions.

## Conclusions

In balanced 1:1 trials, normalizing the FI by total sample size (FQ) or by the arm in which outcomes are toggled (MFQ) differs only by a constant factor (MFQ = 2 × FQ) and therefore conveys identical information about fragility. In trials with unequal allocation, these measures diverge because FI modifies outcomes in a single arm while FQ incorporates both arms in the denominator. In empirical data from large-vessel vasculitis trials and in 93,000 simulation replications, outcome toggles most often occurred in the smaller arm, so total-sample normalization diluted the arm-specific fragility signal by including participants who did not contribute to the statistical reversal. These findings indicate that arm-specific normalization isolates fragility to the population in which outcome changes flip statistical significance, whereas total-sample normalization blends modified and unmodified populations and can mischaracterize trial fragility under unequal allocation.

## Declarations

### Funding

No specific funding received.

### Conflicts of Interest

None declared.

### Data Availability

Simulation code is archived at Zenodo: 10.5281/zenodo.17560282. Empirical data are provided in Supplementary File S2.

### Ethics

Not required (secondary analysis of published data).

### Author Contributions

JJG – writing – original draft, writing –review & editing, project administration; ML – writing – review and editing; DYT – writing – review and editing; TFH-conceptualization, data curation, formal analysis, investigation, methodology, project administration, software, supervision, validation, visualization, writing – original draft, writing – review & editing.

## Supporting information

S1 File. SAMPL Guidelines Checklist.

S2 File. Listing of empiric data and trials analyzed.

## Notes

### Competing Interest Statement

The authors have declared no competing interest.

### Funding Statement

The author(s) received no specific funding for this work.

### Author Declarations

n/a. This research analyzed simulated data and aggregate outcome counts extracted from previously published randomized controlled trials. No individual-level or identifiable participant data were used. Because all data were either synthetically generated or publicly available in aggregate form, this work does not constitute human subjects research and did not require IRB review.

